# Seroconversion rate and socioeconomic and ethnic risk factors for SARS-CoV-2 infection in children in a population-based cohort

**DOI:** 10.1101/2021.10.21.21265322

**Authors:** F Brinkmann, HH Diebner, C Matenar, A Schlegtendal, L Eitner, N Timmesfeld, C Maier, T Lücke

## Abstract

**Introduction:** Socioeconomic and ethnic background have been discussed as possible risk factors for SARS-CoV-2 infections in children. Improved knowledge could lead way to tailored prevention strategies and help to improve infection control.

**Methods:** Observational population-based cohort study in children (6mo. - 18 ys.) scheduled for legally required preventive examination and their parents in a metropolitan region in Germany. Primary endpoint was the SARS-CoV-2 seroconversion rate during study period. Risk factors assessed included age, pre-existing medical conditions, socioeconomic factors, and ethnicity.

**Results:** 2124 children and their parents were included. Seroconversion rates among children in all age groups increased by 3-4-fold from 06/2020 to 02/2021. Only 41% of seropositive children were symptomatic. In 51% of infected children at least one parent was also SARS-CoV-2 positive.

Low level of parental education (OR 3.13 (0.72-13.69)) significantly increased the risk of infection. Of the total cohort, 38.5% had a migration background. Specifically, 9% were of Turkish and 5% of Middle Eastern origin. These children had the highest risk for SARS-CoV-2 infections (OR 6.24 (1.38-28.12) and 6.44 (1.14-36.45) after adjustment for other risk factors.

**Discussion:** Seroprevalence of SARS-CoV-2 infections in children increased by 3-4-fold within the study period. Frequently, more than one family member was infected. Children from families with lower socioeconomic status were at higher risk. The highest risk for SARS-CoV-2 infection was identified in families with Turkish or Middle Eastern background. Culture sensitive approaches are essential to improve infection control and serve as a blueprint for vaccination strategies in this population.

**Trial Registration:** BMBF funding registration 01KI20173 (Corkid)

## Main text

## Introduction

The rate of SARS-CoV-2 infections in children has been discussed controversially during the first months of the pandemic. Children are frequently oligo- or asymptomatic and, therefore, have been undertested and underdiagnosed. Seroconversion studies, however, reveal that the rate of SARS-CoV-2 infections in adults and children are similar [1].

Most infections probably take place in households where contacts are most intense [2]. Cramped living conditions and poverty have been associated with a higher rate of SARS-CoV-2 infection both in adults and children as well as increased morbidity and mortality [3,4]. Ethnic origin has also been described as a risk factor for hospitalization for COVID19 in children in the US [5,6]. Moreover, socioeconomic factors have been identified as risk factors for SARS-CoV-2 infection [3,7].

This study in an unselected asymptomatic paediatric population in a Western German metropolitan region with about 40% of families with migration background had been set up to identify risk factors for SARS-CoV-2 infection in children and adolescents.

## Methods

### Study design and participants

In a standardized approach from 06/2020 to 02/2021 all asymptomatic children and adolescents who attended outpatient paediatric practices in three regions of Western Germany for scheduled mandatory routine examinations (U-Untersuchungen) from 6 months to 18 years of age and their parents were invited to participate in the study. Participants and their parents were asked to fill in a tablet-based questionnaire available in 5 different languages. Questions included former SARS-CoV-2 infections, medical history as well as information on socioeconomic and migration background (Germany/Western Europe, Turkey, Middle East, America, Eastern and Southern Europe, and Asia). Education level was defined as the highest level of education of one of the parents/guardians. High level of education included high school/grammar school (Fachhochschule/Abitur), medium level education general secondary school (Realschule) and low-level education included education up to primary school/basic secondary school (Grundschule/Hauptschule). No educational degree formed a fourth group. Information on chronic diseases and medication was obtained from parents and verified by matching with medical charts.

Serum samples were analysed for SARS-CoV-2 IgM and IgG antibodies (ECLIA, Roche).

The target variable was SARS-CoV-2 infection given by seropositivity or positive PCR test, respectively. We aimed at comparing demographics (age, sex), underlying medical conditions, socioeconomic and migration background between children and adolescents with and without evidence of SARS-CoV-2 infection.

### Statistical Analysis

Descriptive statistics are depicted in tables 1 and 2. Demographic and clinical parameters (table 1) are presented along with univariate odds ratios reported with 95% confidence intervals. In addition, the impact of migration background has been analysed with adjusted random effect logistic regression (cf. table 3). Thereby, we adjusted for the ratio of number of available rooms to the number of persons living in the same household as well as for the highest educational level within the family. The ratio has been introduced to account for the correlation between number of rooms and number of persons following the rational that the available space per person is the essential factor. Within the Corkid study, some families participated with more than one child. Therefore, we included a random effect in our model given by a family ID. Insignificance of other factors has been shown by a likelihood ratio-based model reduction process. For the random effect analysis, the lme4 software package within statistical programming language R has been used [8]. P-values less than 0.05 were considered statistically significant.

**Table 1:** Demographic and clinical data of participants

|  | All | SARS-COV-2<br>positive | SARS-Cov-2 nega-<br>tive | odds ratio<br>(SARS<br>yes/no) |
| --- | --- | --- | --- | --- |

| **N** | **2124** | **58** | **2066** |  |
| --- | --- | --- | --- | --- |
| **Girls** | 1020 (48.02%) | 27 (46.55%) | 993 (48.06%) | 0.94 (0.56-1.59) |
| **<= 3 years** | 429 (20.2%) | 13 (22.41%) | 416 (20.14%) | 1.15 (0.61-2.14) |
| **4-6 years (pre-school)** | 817 (38.47%) | 22 (37.93%) | 795 (38.48%) | 0.98 (0.57-1.67) |
| **7-10 years (primary school)** | 290 (13.65%) | 8 (13.79%) | 282 (13.65%) | 1.01 (0.47-2.16) |
| **11-18 years (secondary school)** | 588 (27.68%) | 15 (25.86%) | 573 (27.73%) | 0.91 (0.5-1.65) |
| **Underweight (<3 percentile)** | 159 (7.49%) | 3 (5.17%) | 156 (7.55%) | 0.67 (0.21-2.16) |
| **Obesity (>97% percentile)** | 156 (7.34%) | 4 (6.9%) | 152 (7.36%) | 0.93 (0.33-2.61) |
| **Any chronic medical condition** | 389 (18.31%) | 8 (13.79%) | 381 (18.44%) | 0.71 (0.33-1.5) |
| **Chronic Airway Disease** | 211 (9.93%) | 4 (6.9%) | 207 (10.02%) | 0.67 (0.24-1.86) |

**Table 2a:** Socioeconomic and migration background of participants

|  | **All** | **SARS-CoV-2**<br>**positive** | **SARS- CoV-2**<br>**negative** |
| --- | --- | --- | --- |
| **N (all=100%)** | 2124 | 58 | 2066 |
| **Parents SARS-CoV-2 positive** | 61 (3.07%) | 28 (51.85%) | 33 (1.71%) |
| **SARS-CoV-2 Contact** | 261 (13.12%) | 32 (59.26%) | 229 (11.83%) |
| **Parents in high-risk occupations for Sars-CoV-2** | 995 (50.03%) | 27 (50%) | 968 (50.03%) |
| **Parents Highest Educational Degree** |  |  |  |
| **High School/Grammar School** | 1311 (65.91%) | 12 (22.22%) | 629 (32.51%) |
| **General Secondary School** | 240 (12.07%) | 7 (12.96%) | 233 (12.04%) |
| **Basic Secondary /Primary School** | 66 (3.32%) | 6 (11.11%) | 60 (3.1%) |
| **No educational degree** | 70 (5.56%) | 3 (6.25%) | 67 (3.46%) |
| **Migration Background Parents/Grandparents** | 766 (38.51%) | 35 (64.81%) | 731 (37.78%) |
| **Turkey** | 180 (9.05%) | 13 (24.07%) | 167 (8.63%) |
| **Middle East** | 108 (5.43%) | 8 (14.81%) | 100 (5.17%) |
| **Asia** | 28 (1.41%) | 3 (5.56%) | 25 (1.29%) |
| **CIS States (former Soviet Union)** | 127 (6.39%) | 3 (5.56%) | 124 (6.41%) |
| **Amerika** | 11 (0.55%) | 0 (0%) | 11 (0.57%) |
| **Eastern Europe** | 288 (14.48%) | 7 (12.96%) | 281 (14.52%) |
| **Southern Europe** | 51 (2.56%) | 1 (1.85%) | 50 (2.58%) |
| **Germany/Western Europe** | 1257 (63.2%) | 20 (37.04%) | 1237 (63.93%) |
| **Members Household** |  |  |  |
| **1-3** | 529 (24.91%) | 11 (18.97%) | 518 (25.07%) |
| **4-5** | 1248 (58.76%) | 35 (60.34%) | 1213 (58.71%) |
| **> 5** | 156 (7.34%) | 7 (12.07%) | 149 (7.21%) |
| **Rooms/Apartment** |  |  |  |
| **1-3** | 456 (22.93%) | 21 (38.89%) | 435 (22.48%) |
| **4-5** | 967 (48.62%) | 21 (38.89%) | 946 (8.89%) |
| **>6** | 504 (25.34%) | 11 (20.37%) | 493 (25.48%) |

**Table 2b:** Subgroup analysis of migration background and number of rooms/persons

| **Predictors** | **Total** | **SARs-Cov-2 positive** | **SARS- CoV-2 negative** | **Odds ratio/ 95% CI** |
| --- | --- | --- | --- | --- |
| **< 1 room/person & no migration background** | 129 (6,49%) | 5 (9,26%) | 124 (6,41%) | 1,48 (0,58-3,76) |
| **>= 1 room/person & no migration background** | 1037 (52,14%) | 13 (24,07%) | 1024 (52,92%) | 0,29 (0,16-0,55) |
| **< 1 room/person & migration background** | 270 (13,57%) | 17 (31,48%) | 253 (13,07%) | 2,97 (1,66-5,31) |
| **>= 1 room/person & migration background** | 488 (24,53%) | 18 (33,33%) | 470 (24,29%) | 1,53 (0,87-2,69) |
| **< 1 room/person & Turkish background** | 69 (3,47%) | 8 (14,81%) | 61 (3,15%) | 5,26 (2,39-11,57) |
| **>= 1 room/person & Turkish background** | 97 (4,88%) | 5 (9,26%) | 92 (4,75%) | 2,02 (0,79-5,18) |
| **< 1 room/person & Middle Eastern European background** | 68 (3,42%) | 4 (7,41%) | 64 (3,31%) | 2,32 (0,81-6,59) |
| **>= 1 room/person & Middle Eastern background** | 35 (1,76%) | 4 (7,41%) | 31 (1,6%) | 4,86 (1,66-14,26) |
| **< 1 room/person & Eastern European background** | 79 (3,97%) | 3 (5,56%) | 76 (3,93%) | 1,43 (0,44-4,67) |
| **>= 1 room/person & Eastern European background** | 185 (9,3%) | 3 (5,56%) | 182 (9,41%) | 0,56 (0,17-1,82) |

**Table 3:** Results of random effect logistic regression. Intra class correlation amounts to ICC=0.7.

| Predictors | OR (logistic regression) | 95% -CI | p- value | OR (univariate) | p-value |
| --- | --- | --- | --- | --- | --- |
| **Background** |  |  |  |  |  |
| **Germany/Western Europe** | 1.0 | - | - | - | - |
| **Turkey** | 6.24 | 1.38-28.13 | 0.017 | 4.81 | < 0.001 |
| **Middle East** | 6.44 | 1.14- 36.45 | 0.035 | 4.95 | < 0.001 |
| **Eastern Europe** | 1.56 | 0.42-5.78 | 0.503 | 1.54 | n.s. |
| **Asia** | 16.36 | 1.07- 250.36 | 0.045 | 7.42 | 0.002 |
| **Other** | 1.26 | 0.4-11.28 | 0.835 | 1.5 | n.s. |
| **Living conditions** |  |  |  |  |  |
| **Rooms/person** | 0.35 | 0.08-1.5 | 0.158 | 0.25 | 0.001 |
| **Level of education** |  |  |  |  |  |
| **Higher level** | 1.0 | - | - | - | - |
| **Medium level** | 1.02 | 0.37-2.81 | 0.972 | 1.08 | Ns |
| **Lower level** | 3.13 | 0.72-13.69 | 0.129 | 3.15 | 0.003 |
| **No educational degree** | 1.63 | 0.23-11.69 | 0.628 | 1.6 | Ns |

## Results

### Demographics and Clinical Characteristics

In this cohort, initially 2848 children and adolescents were asked to participate, 25% refused, mainly because of fear of drawing blood.

Of the remaining 2124 (1019 girls; 48%) with medium age 7.1 years [0.6-18], 58 (2.78%) had evidence of SARS-CoV-2 infection. There were no clinical differences to SARS-CoV-2 negative children. Also, the number of children with pre-existing conditions (13% vs 18%) did not significantly differ between the groups. 24 of the 58 infected children (41.3%) did not recall any infection during the preceding three months.

Clinical characteristics of the participants are displayed in table 1.

### Socioeconomic and migration background

Within the study cohort, 766 (38.5%) children and adolescents had a migration background see table 2). Whereas 4.5% of children and adolescents with a migration background had evidence of SARS-CoV-2 infection, only 1.6% of children with Western European background had evidence of infection (OR 2.78 [1.63-4.74]). Details of socioeconomic and migration background of participants are shown in table 2a and table 2b.

A random effect logistic regression yields the results listed in table 3. Specifically, the highest risks for infection were detected in children of Turkish (OR 6.24 [1.38-28.13], p=0.017) and Middle Eastern (OR 6.44 [1.14-36.45], p=0.035) origin, respectively.

In the Asian population seroprevalence was also high (OR 16.6 [1.07-250]), but because of the small number of participants (1.4% of the cohort) from this region of origin this result must be interpreted with caution.

Other risk factors relevant for risk adjustment, although below significance, included low level education of the parents (OR 3.13 [0.72-13.69]) and crowded living conditions inversely expressed by rooms per person (OR 0.35 [0.72-13.69]). Of note, these two factors turn out to have statistically significant effects in univariate analyses (see table 3).

## Discussion

This large population-based study identifies seroconversion rates and risk factors for SARS-CoV-2 infections in asymptomatic children and adolescents, attended legally required preventive examinations. Our key findings are:

- less than half of seropositive participants of all age groups did recall symptoms of infection suggesting that more than half of SARS-CoV-2 infections go unnoticed [9].
- the incidence of SARS-CoV-2 infections is significantly higher in families of Turkish or Middle Eastern decent independent of other risk factors i.e. after adjustment for other socioeconomic confounders.

Our cohort had increasing seroprevalence rates from 0.5% in Mid-2020 to almost 6% in the beginning of 2021 like other seroprevalence data from German or Swiss school cohorts [10,11]. The dynamics in increase of seroprevalence of SARS-CoV-2 infections matches well with national data of acute SARS-CoV-2 infection in the same geographic region [2,12]. As shown before [2] infection rates have been comparable across all age groups in keeping with the findings of other authors [1,6].

It is not surprising, that the known exposure to SARS-CoV-2 positive individuals doubled the risk for seroconversion. In most families, at least one parent also has evidence of SARS-CoV-2 infection, which is slightly more than described in other cohorts [13,14]. However, 50% of these SARS-CoV-2 exposed children did not develop evidence of infection.

Recent study from the US state that socioeconomically disadvantaged children, especially those from ethnic minorities, are at higher risk of infection [3,5,6]. Adult data from the UK and the US also show increased morbidity and mortality from SARS-CoV-2 in adults from ethnic minorities and with poor socioeconomic status. [4,7]. Limited access to healthcare systems and migration background might also play a role in spreading the disease [15,16].

The most relevant risk factor for SARS-CoV-2 infection in our study population is a Turkish or Middle Eastern migration background. Although socioeconomic factors confound the factor of migration history to some extent, our findings result from an adjusted regression model and are therefore independent of educational standard and housing conditions.

Reasons for this observation could include different family and social structures favouring closer contacts and therefore increasing the risk of transmission as described in a tight knit Jewish Orthodox community in the US by Gaskell et al. [17]. Apart from that, language as well as cultural barriers might play a role [7]. Visiting friends and relatives in countries with higher incidence of infection might also further increase the risk. Scepticism regarding politics and health authorities are an additional issue, especially in prevention and vaccination programs [1]. To approach these families, tailored, culture sensitive strategies need to be developed and can serve as a blueprint for vaccination programs.

Limitations: Paediatric patients were recruited only from three regions (counties) in Western Germany and not all paediatric practices in the area participated. In addition, initially low seroprevalence rates could have introduced a bias. Genetic risk factors could predispose certain populations to infections with SARS-CoV-2, which was not considered in this study. The proportion of children from Turkish families, in contrast to those of other origins, was lower than expected at 9% (see table 1), whereas ca. 20% of the families in the regions studied are of Turkish origin. However, this rather indicates an underestimation of the true infection rate in this group. Another limitation might be the use of surrogate parameters for the assessment of living conditions and income like number of rooms etc. Different educational grades might have posed difficulties in answering questions about educational status and might have also led to an underestimation of the corresponding risk factor. We attempted to reduce a possible bias through semi structured interviews to validate the answers drawn from the internet-based questionnaire.

## Conclusions

This study shows that poor socioeconomic status and Turkish or Middle Eastern migration background are independent risk factors for SARS-CoV-2 infection in a population-based cohort of children and adolescents in Germany.

Targeted interventions and tailored prevention strategies in these groups are necessary to improve infection control and to protect these vulnerable populations from excess morbidity and mortality. They could serve as a blueprint for vaccination programs and there is need to shift the focus of politics accordingly.

[Back to top](https://jamanetwork.com/journals/jamapediatrics/pages/instructions-for-authors#top)

## Data Availability

All data produced in the present study are available upon reasonable request to the authors

## Notes

### Competing Interest Statement

The authors have declared no competing interest.

### Funding Statement

This study was funded by BMBF: funding registration 01KI20173 (Corkid)

### Author Declarations

The Ethics committee of Ruhr University Bochum gave ethical approval for this work

